# Postoperative 20% Albumin and Cardiac Surgery Associated Kidney Injury, Statistical Analysis Plan and Updated Protocol

**DOI:** 10.1101/2024.09.17.24313807

**Authors:** Mayurathan Balachandran, Adrian Pakavakis, Wisam Al-Bassam, David Collins, Raffaele Mandarano, Vineet Sarode, Rinaldo Bellomo, Alastair Brown, Shailesh Bihari, Mozhu Li, Alana Brown, Yahya Shehabi

## Abstract

**Background:** The incidence of cardiac surgery associated acute kidney injury (CS-AKI) remains high. Patients who develop AKI after cardiac surgery are at higher risk of persistent renal dysfunction and increased long-term mortality. The risk of CS-AKI is significantly increased in patients with chronic kidney disease and in patients having prolonged bypass for complex surgery. Previous trials of albumin did not show any benefit in prevention of CS-AKI. These trials, however, did not focus on high-risk patients and used albumin as a resuscitation strategy. The aim of ALBICS-AKI is to demonstrate the effect of concentrated albumin infusion on CS-AKI in high-risk patients undergoing cardiac surgery compared with standard care.

**Methods:** ALBICS-AKI is an investigator initiated, multicentre, randomised, open label trial. Seven centres in Australia and Italy will participate in the trial. We will randomise 620 adult patients who will undergo on-pump cardiac surgery with one of the following: an estimated glomerular filtration rate <60 ml/min/1.73m^2^, combined valve/s, coronary artery, or surgery involving thoracic aorta. Within 6 hours after surgery, a 20% albumin infusion will commence at 20ml/h for 15 hours. All patients will receive standard care as per institutional protocols. The primary outcome is the proportion of patients with AKI according to creatinine based KDIGO definition at hospital discharge or day 28, whichever comes first. Secondary outcomes include Major Adverse Kidney Events at day 28, AKI stage II and III, need for renal replacement therapy, and hospital mortality.

**Ethics and dissemination:** The trial was approved by Monash Health Lead Research Committee for Australian sites and by the Italian Medicine Agency for Italian sites. The estimated study completion date is Sep 2024. The results will be presented at major conferences and submitted for publication in peer-reviewed journals.

**Trial registration number:** Australian New Zealand Clinical Trials Registry ACTRN12619001355167

## INTRODUCTION

Cardiac surgery associated acute kidney injury (CS-AKI) is common. In a cohort of more than 25,000 patients, the reported incidence of CS-AKI was 30% [1]. Recent trials investigating CS-AKI in high-risk populations, based on factors such as surgical complexity and baseline renal function, suggested a background incidence as high as 74% [2–5]. Persistent renal dysfunction at 90-days occurred in 24% of high-risk patients who developed CS-AKI [6] and was associated with increased long-term mortality and risk of severe chronic kidney disease [7–9]. The burden of CS-AKI is likely to increase with increasing comorbid conditions, and complexity of cardiac surgery [10,11]. With more than one million patients undergoing cardiac surgery every year, prevention of CS-AKI remains a priority [12].

The role of perioperative albumin administration in cardiac surgery remains controversial. In off-pump surgery, a trial of preoperative 20% albumin reported a reduction in CS-AKI [13]. Albeit with some limitations, a single centre trial investigating the use of perioperative 4% albumin infusion did not significantly reduce CS-AKI or major adverse events after cardiac surgery [14]. Notably this study was limited by the inclusion of low-risk patients with just 3% of patients suffering AKI, well below estimates in the literature. Further, mean baseline GFR was 80ml/min/1.73m^2^, mean EUROScore 1.7, and 81% of patients underwent only a single cardiac procedure.

A phase II multicentre trial (HAS FLAIR-II) of 20% Human Albumin Solution Fluid Bolus Administration Therapy in Patients after Cardiac Surgery-II (HAS FLAIR-II), to determine whether fluid resuscitation with 20% albumin (Albumex®20) decreases the duration of vasopressor requirement compared to usual care, has been completed. This trial, however, was not powered to detect CS-AKI [15].

Although without strong evidence, a recent consensus statement and systematic review did not recommend the use of albumin as a first line fluid in priming or resuscitation in cardiac surgery and called for more investigation in this area [16].

Our first study protocol was published in 2021[17]. Recruitment was halted completely during the COVID pandemic shortly after commencement in 2020 till late 2022 (Figure 1). Since then, we have new sites joining the trial in Australia and Italy. In addition, the composition of normal serum albumin changed from 4% to 5% in late 2023. Some centres use normal serum albumin as a resuscitative fluid in standard care. At the same time, the composition of 20% albumin changed with increased osmolality and sodium content.

**Figure 1.**
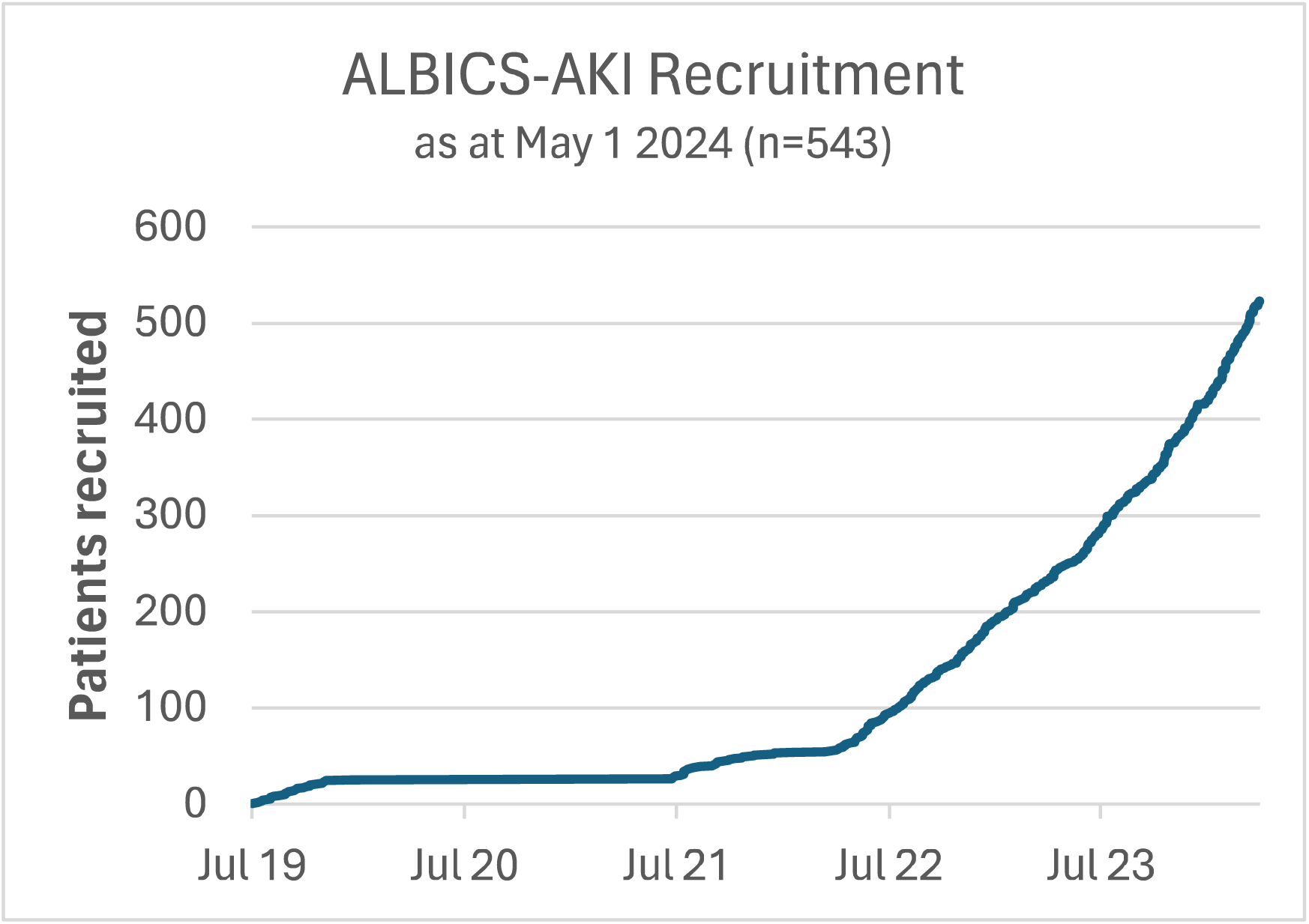
Recruitment progress over time, up to May 1 2024. Recruitment was halted during the COVID-19 pandemic between 2019 and 2021. More than 90% of patients were recruited after July 2022.

We present a detailed statistical analysis plan and an updated protocol. The changes in the updated protocol and rationale for these changes are highlighted in Table 1.

**Table 1.**
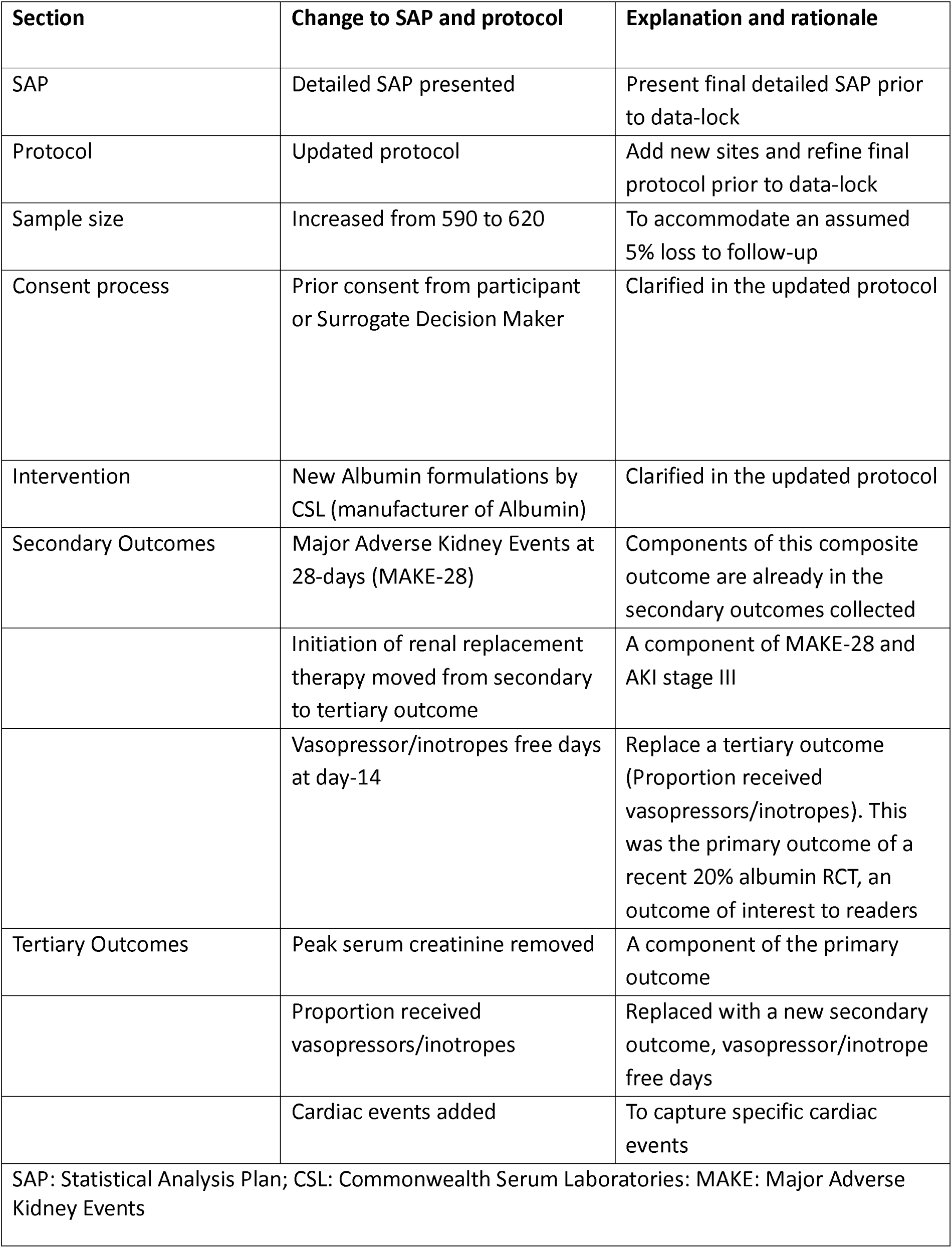
Changes made in the detailed statistical analysis plan and updated protocol and their rationale.

**Table 2.**
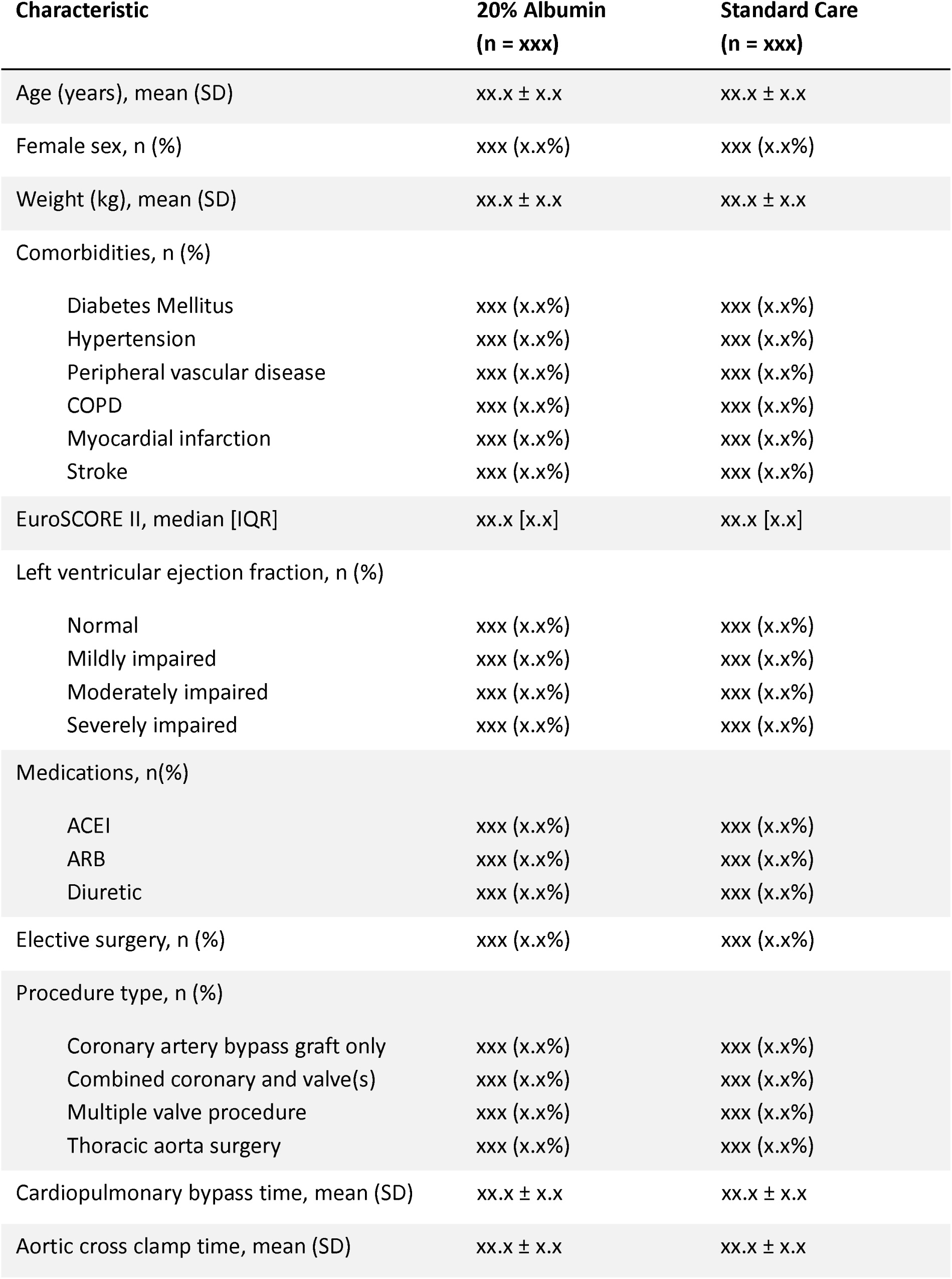
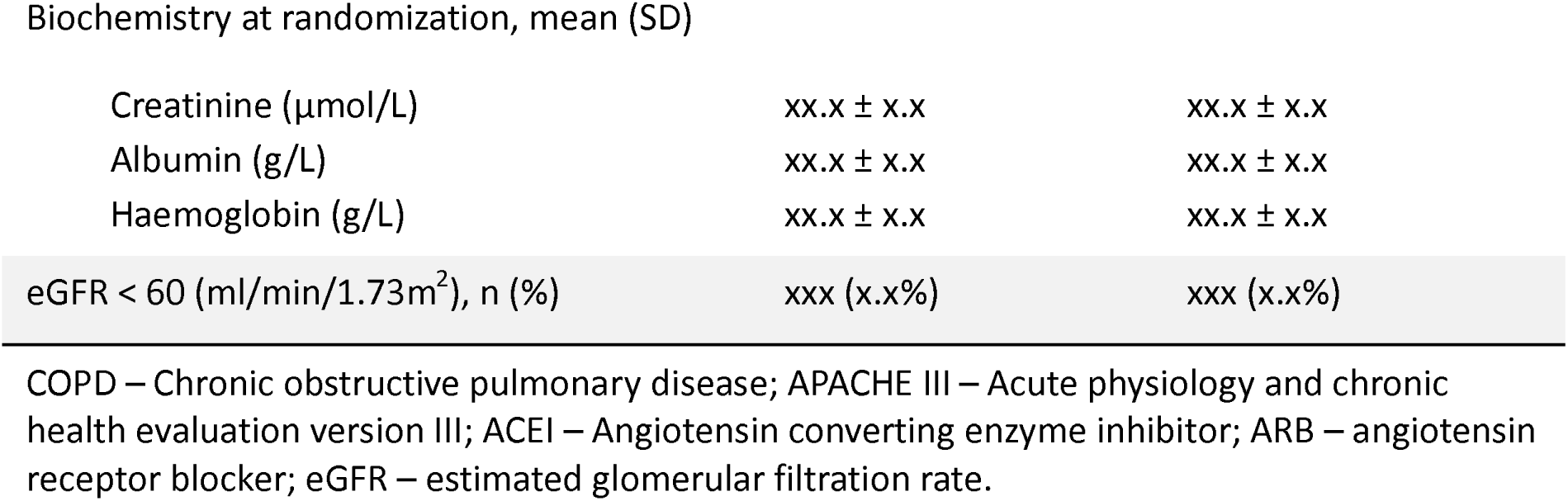
Baseline characteristics by randomisation group.

| Characteristic | 20% Albumin<br>(n = xxx) | Standard Care<br>(n = xxx) |
| --- | --- | --- |
| Age (years), mean (SD) | xx.x ± x.x | xx.x ± x.x |
| Female sex, n (%) | xxx (x.x%) | xxx (x.x%) |
| Weight (kg), mean (SD) | xx.x ± x.x | xx.x ± x.x |
| Comorbidities, n (%) |  |  |
| Diabetes Mellitus | xxx (x.x%) | xxx (x.x%) |
| Hypertension | xxx (x.x%) | xxx (x.x%) |
| Peripheral vascular disease | xxx (x.x%) | xxx (x.x%) |
| COPD | xxx (x.x%) | xxx (x.x%) |
| Myocardial infarction | xxx (x.x%) | xxx (x.x%) |
| Stroke | xxx (x.x%) | xxx (x.x%) |
| EuroSCORE II, median [IQR] | xx.x [x.x] | xx.x [x.x] |
| Left ventricular ejection fraction, n (%) |  |  |
| Normal | xxx (x.x%) | xxx (x.x%) |
| Mildly impaired | xxx (x.x%) | xxx (x.x%) |
| Moderately impaired | xxx (x.x%) | xxx (x.x%) |
| Severely impaired | xxx (x.x%) | xxx (x.x%) |
| Medications, n(%) |  |  |
| ACEI | xxx (x.x%) | xxx (x.x%) |
| ARB | xxx (x.x%) | xxx (x.x%) |
| Diuretic | xxx (x.x%) | xxx (x.x%) |
| Elective surgery, n (%) | xxx (x.x%) | xxx (x.x%) |
| Procedure type, n (%) |  |  |
| Coronary artery bypass graft only | xxx (x.x%) | xxx (x.x%) |
| Combined coronary and valve(s) | xxx (x.x%) | xxx (x.x%) |
| Multiple valve procedure | xxx (x.x%) | xxx (x.x%) |
| Thoracic aorta surgery | xxx (x.x%) | xxx (x.x%) |
| Cardiopulmonary bypass time, mean (SD) | xx.x ± x.x | xx.x ± x.x |
| Aortic cross clamp time, mean (SD) | xx.x ± x.x | xx.x ± x.x |
| Biochemistry at randomization, mean (SD) |  |  |
| Creatinine (μmol/L) | xx.x ± x.x | xx.x ± x.x |
| Albumin (g/L) | xx.x ± x.x | xx.x ± x.x |
| Haemoglobin (g/L) | xx.x ± x.x | xx.x ± x.x |
| eGFR < 60 (ml/min/1.73m<sup>2</sup>), n (%) | xxx (x.x%) | xxx (x.x%) |

COPD – Chronic obstructive pulmonary disease; APACHE III – Acute physiology and chronic health evaluation version III; ACEI – Angiotensin converting enzyme inhibitor; ARB – angiotensin receptor blocker; eGFR – estimated glomerular filtration rate.

**Table 3.** ICU interventions by randomization group Intervention 20% Albumin.

| **Intervention** | **20% Albumin**<br>**(n = xxx)** | **Standard Care**<br>**(n = xxx)** | **P-value** |
| --- | --- | --- | --- |
| **Cardiovascular** |  |  |  |
| Proportion of patients who received inotropes or vasopressors on: |  |  |  |
| Day 1, n (%) | xxx (x.x%) | xxx (x.x%) | xxx |
| Day 2, n (%) | xxx (x.x%) | xxx (x.x%) | xxx |
| Day 3, n (%) | xxx (x.x%) | xxx (x.x%) | xxx |
| Day 4, n (%) | xxx (x.x%) | xxx (x.x%) | xxx |
| Day 5, n (%) | xxx (x.x%) | xxx (x.x%) | xxx |
| Day 6, n (%) | xxx (x.x%) | xxx (x.x%) | xxx |
| Day 7, n (%) | xxx (x.x%) | xxx (x.x%) | xxx |
| After day 7, n (%) | xxx (x.x%) | xxx (x.x%) | xxx |
| IABP insertion, n (%) | xxx (x.x%) | xxx (x.x%) | xxx |
| **Fluids administered by the end of Day 2** |  |  |  |
| Normal serum Albumin (ml), mean (SD) | xx.x ± x.x | xx.x ± x.x | xxx |
| n (%)patients | xxx (x.x%) | xxx (x.x%) | xxx |
| 20% Albumin (ml), mean (SD) | xx.x ± x.x | xx.x ± x.x | xxx |
| n (%) patients | xxx (x.x%) | xxx (x.x%) | xxx |
| Crystalloid (ml), mean (SD) | xx.x ± x.x | xx.x ± x.x | xxx |
| n (%) patients | xxx (x.x%) | xxx (x.x%) | xxx |
| Packed red cells (ml), mean (SD) | xx.x ± x.x | xx.x ± x.x | xxx |
| n (%)patients | xxx (x.x%) | xxx (x.x%) | xxx |
| Other blood products (ml), mean (SD) | xx.x ± x.x | xx.x ± x.x | xxx |
| n (%) patients | xxx (x.x%) | xxx (x.x%) | xxx |
| **Diuretic therapy** |  |  |  |
| Intravenous loop, n (%) | xxx (x.x%) | xxx (x.x%) | xxx |
| Other class, n (%) | xxx (x.x%) | xxx (x.x%) | xxx |

IABP – Intra aortic balloon pump

**Table 4.** Primary, secondary and tertiary outcomes.

|  | 20% Albumin<br>(n = xxx) | Standard Care<br>(n = xxx) | Hazard Ratio<br>Estimate<br>(95% CI) | Adjusted Hazard<br>ratio estimate<br>(95% CI) | p |
| --- | --- | --- | --- | --- | --- |
| **Primary outcome** |  |  |  |  |  |
| In-hospital AKI at Day 28, n (%) | xxx (x.x%) | xxx (x.x%) | xx (xx-xx) | xx (xx-xx) | x.xx |
| eGFR > 60 (ml/min/1.73m<sup>2</sup>) | xxx (x.x%) | xxx (x.x%) | xx (xx-xx) | xx (xx-xx) | x.xx |
| eGFR < 60 | xxx (x.x%) | xxx (x.x%) | xx (xx-xx) | xx (xx-xx) | x.xx |
| **Secondary outcomes** |  |  |  |  |  |
| MAKE at day 28, n (%) | xxx (x.x%) | xxx (x.x%) | xx (xx-xx) | xx (xx-xx) | x.xx |
| eGFR > 60 (ml/min/1.73m<sup>2</sup>) | xxx (x.x%) | xxx (x.x%) | xx (xx-xx) | xx (xx-xx) | x.xx |
| eGFR < 60 | xxx (x.x%) | xxx (x.x%) | xx (xx-xx) | xx (xx-xx) | x.xx |
| Hospital mortality Day 28, n (%) | xxx (x.x%) | xxx (x.x%) | xx (xx-xx) | xx (xx-xx) | x.xx |
| AKI stage II or III, Day 28, n (%) | xxx (x.x%) | xxx (x.x%) | xx (xx-xx) | xx (xx-xx) | x.xx |
| CRRT by Day 28 or Hospital discharge | xxx (x.x%) | xxx (x.x%) | xx (xx-xx) | xx (xx-xx) | x.xx |
| Length of ICU stay (hours), median (IQR) | xx.x ± x.x | xx.x ± x.x | xx (xx-xx) | xx (xx-xx) | x.xx |
| Length of hospital days (days), median (IQR) | xx.x ± x.x | xx.x ± x.x | xx (xx-xx) | xx (xx-xx) | x.xx |
| Duration of mechanical ventilation (hours), median (IQR) | xx.x ± x.x | xx.x ± x.x | xx (xx-xx) | xx (xx-xx) | x.xx |
| Vasopressor/inotrope free days at Day 14, median [IQR] | xx.x [x.x] | xx.x [x.x] | xx (xx-xx) | xx (xx-xx) | x.xx |
| **Tertiary outcomes** |  |  |  |  |  |
| Serum albumin at 24 hours (g/L), mean (SD) | xx.x ± x.x | xx.x ± x.x | xx (xx-xx) | xx (xx-xx) | x.xx |
| Net fluid balance at end of Day 2 (ml), median (IQR) | xx.x ± x.x | xx.x ± x.x | xx (xx-xx) | xx (xx-xx) | x.xx |
| Blood transfusion requirements at end of Day 2 (ml), median (IQR) | xx.x ± x.x | xx.x ± x.x | xx (xx-xx) | xx (xx-xx) | x.xx |
| Life-threatening arrhythmia, n (%) | xxx (x.x%) | xxx (x.x%) | xx (xx-xx) | xx (xx-xx) | x.xx |

AKI – Acute kidney injury; MAKE – Major adverse kidney events; eGFR – estimated glomerular filtration rate; ICU – intensive care unit; IQR – Inter Quartile Range

## MATERIALS AND METHODS

### Trial design

The ALBICS-AKI trial is a pragmatic multi-centre, parallel-group, open label, prospective randomised controlled superiority trial. Patients will be randomised in a 1:1 ratio to receive either an infusion of 20% albumin or standard care. Patients are stratified at randomisation according to study site and preoperative eGFR above or below 60ml/min/1.73m^2^.

Our hypothesis is that a postoperative continuous infusion of 20% albumin reduces the incidence of cardiac surgery associated acute kidney injury (CS-AKI) in high-risk patients.

### Selection of patients

#### Inclusion criteria

- Aged 18 years or older at the time of screening
- Have undergone cardiac or thoracic aortic surgery
- At least one of the following:
  - Estimated glomerular filtration rate (eGFR) < 60ml/min/1.73m^2^
  - Undergoing a combined valve and coronary artery procedure
  - Undergoing two or more valve procedures
  - Surgery involving the thoracic aorta Exclusion criteria
- eGFR < 15ml/min/1.73m^2^
- Serum albumin < 20g/L
- Dialysis dependence
- Kidney transplant recipient
- Undergone off-pump cardiac surgery
- Patient been in intensive care more than 6 hours after surgery
- Requiring extra-corporeal life support or ventricular assist device immediately postoperative
- Jehovah’s Witness

### Recruitment modality and consent

Patients meeting all inclusion criteria will be screened for eligibility. Whenever possible, consent will be obtained from participants prior to surgery. If this is not feasible, consent will be sought from the participant or their surrogate decision-maker soon after arrival to intensive care and before randomization. Once extubated and practical, participants will provide their own consent to continue in the trial. Patient flow through the trial will be reported in a Consolidated Standards of Reporting Trials (CONSORT) diagram (Figure 2) [18]. The diagram will summarise the numbers of patients screened, eligible, randomised and analysed. Reasons for ineligibility, inability to randomise or analyse will be summarised in the diagram.

**Figure 2.**
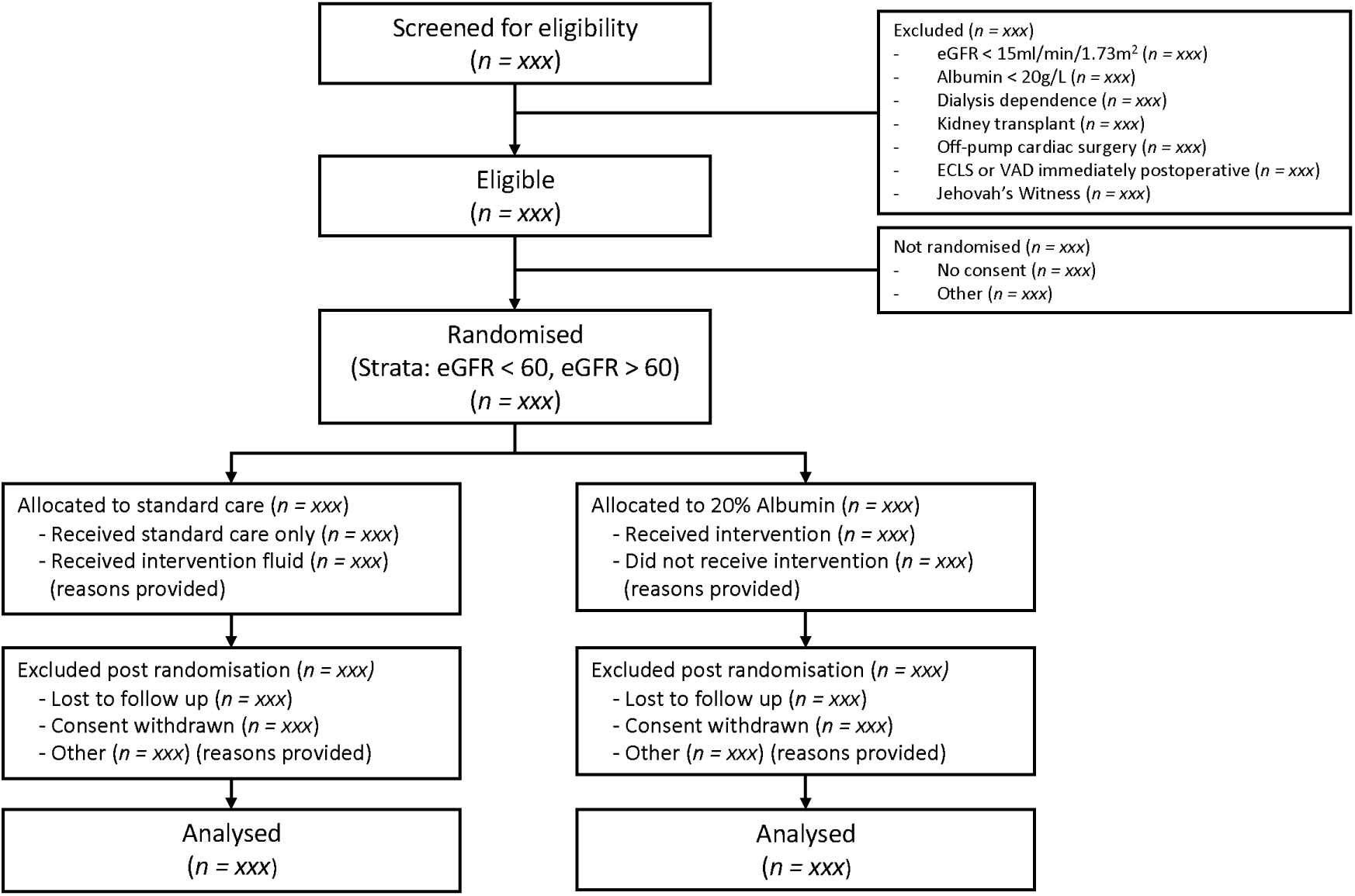
Patient flow diagram. eGFR – estimated glomerular filtration rate; ECLS – extra-corporeal life support; VAD – ventricular assist device.

### Randomisation

A permuted block, computer-generated, randomisation sequence with fixed block size, stratified by hospital and eGFR in 1:1 ratio was generated. Randomisation is conducted via secured Monash University REDCap (Research Electronic Data Capture) [19,20] using a randomisation sequence prepared by the study statistician. Blinding is deemed not feasible. As such, this is an open label trial [17].

### Trial intervention

Patients randomised to the intervention arm will receive a continuous infusion of 20% albumin for 15 hours at 20ml/h commencing within a maximum of 6 hours from ICU admission.

Patients randomised to the standard care arm will not receive 20% albumin for the first 24 hours of ICU admission.

Patients in either arm may receive any concomitant treatment considered routine care at each institution, such as inotropes, vasopressors, diuretic therapy, blood transfusion and normal serum albumin for resuscitation only. Initiation of renal replacement therapy and weaning from ventilation is at the discretion of the treating physician.

### Summary of outcomes

#### Primary outcome

The primary outcome will be in-hospital acute kidney injury of any stage, from the time of randomisation to day 28 or hospital discharge, whichever comes first. Acute kidney injury will be defined using the serum creatinine criteria of the Kidney Disease Improving Global Outcome (KDIGO) definition [21].

#### Secondary Outcomes

Secondary outcomes for the trial will be:

- Major Adverse Kidney Events (MAKE) at day 28. This is a composite outcome of KDIGO defined in-hospital AKI stages II (>100% rise in creatinine) and III (>200% rise in creatinine), requirement for renal replacement therapy, and mortality.
- Hospital mortality to day 28 post ICU admission
- AKI stage II or III, assessed to day 28 or hospital discharge, whichever comes first
- Initiation of continuous renal replacement therapy (CRRT), assessed to day 28 or hospital discharge, whichever comes first
- Length of ICU and hospital stay of the index admission.
- Duration of mechanical ventilation during the index ICU admission.
- Vasopressor/inotrope free days at day 14.

#### Tertiary outcomes

The tertiary outcomes will be:

- Serum albumin level at 24 hours post randomisation
- Net fluid balance at end of post operative day 2
- Blood transfusion requirements at end of post operative day 2
- Occurrence of life-threatening arrhythmia (results in significant haemodynamic compromise, such as supraventricular tachycardia, ventricular tachycardia or fibrillation, atrial fibrillation, sinus pause, and cardiac arrest).

### Patient and public involvement

We acknowledge the benefits of patients/public advocates in the design and conduct of clinical trials. This trial’s timeline, however, did not permit direct consumer advocate engagement from the outset. Thus, while patients/advocates were not directly involved in the design of this study, careful consideration was given to choosing outcomes that are patient centred, clinically meaningful, and relevant. The study team has access to institutional consumer advocates which will be engaged as appropriate following study completion. Patient friendly media summarising relevant findings will be prepared in consultation with consumer advocates and widely disseminated to maximise public understanding of the meaning of the trial.

## STATISTICAL ANALYSIS

This statistical analysis plan has been written in accordance with the ‘Guidelines for the Content of Statistical Analysis Plans in Clinical Trials’ [22].

### Timing of final analysis

The final analysis of all outcomes will occur collectively after the final patient completes the follow up period. All outcomes will be published in the main report. There are no planned interim analyses.

### Timing of outcome assessments

The timeline for outcome assessments is given in the published protocol [17]. The trial intervention will run for a total of 15 hours. All outcomes requiring analysis of blood samples will be taken from routinely collected data, therefore the timing of testing is at the discretion of the treating clinicians.

### Sample size

The incidence of CS-AKI in the standard care arm is expected to be 30%. This baseline rate has been taken from a published cohort study of over 25,000 patients receiving cardiac surgery [23]. A sample size of 590 patients will provide 80% power to detect an absolute reduction of 10% in the primary outcome (incidence of in-hospital CS-AKI up to day 28). Assuming a 5% rate of withdrawal or loss-to-follow up, sample size has been increased to 620 patients.

### Strengths and limitations

The ALBICS-AKI trial aims to differentiate itself from earlier trials in several key areas. First, this trial will investigate the effect of exogenous albumin exclusively in high-risk patients undergoing cardiac surgery. Such patients are at higher risk of CS-AKI and have the potential to see greater benefit from an effective treatment [24–27]. The use of concentrated rather than iso-oncotic albumin preparations seek to leverage albumin’s pharmacological properties in addition to its haemodynamic effects. Further, the deleterious effects of inflammation, oxidative stress, and endothelial dysfunction associated with cardiopulmonary bypass are known to persist into the postoperative period [28,29]. Albumin infusion in the postoperative period, compared to pre- or intraoperative infusion in earlier trials, may offer more benefit in mitigating these effects. Finally, the multi-centre design will strengthen the generalisability of results.

This trial is open label. As such, knowledge of the allocated study arm may impact the treating clinician’s choice of additional therapies. Furthermore, normal serum albumin administration is standard practice in some participating sites. As such it was permitted in the control arm of this trial, which may mask the effect of the study intervention.

## STATISTICAL PRINCIPLES

### Analysis populations

All outcomes will be analysed using an intention-to-treat population for the primary analysis. This population will include all patients according to their allocated treatment arm. Patients who were ineligible, or for whom consent was withdrawn will be excluded. A per-protocol sensitivity analysis will be performed should greater than 5% of the study population have a major protocol deviation, defined as not receiving 20% albumin in the intervention group or receiving 20% albumin in the standard care group in the first 24h after randomisation.

### Confidence intervals and p-Values

All p-values will be reported as two sided with a value less than 0.05 used to determine statistical significance. Confidence intervals will be presented as 95% intervals.

Given the study has a single primary outcome measured only at study completion, adjustment for multiplicity is not required. However, adjustment for multiplicity will be considered for secondary outcomes only, to calculate p-values. All other comparisons, including subgroup analyses of the primary outcome, and sensitivity analyses will be treated as exploratory only and therefore do not require adjustment [30].

### Adherence and protocol deviations

The following pre-specified protocol deviations will be recorded. The total number (%) of each type of deviation will be summarised in the main report:

- Ineligible patient randomised
- Patient randomised to 20% albumin but did not receive the study fluid
- Patient randomised to standard care but received 20% albumin within the first 24 hours

### Baseline characteristics and intervention data

Baseline patient characteristics at the time of randomisation will be summarised by allocation as shown in Table. The table will show basic physical characteristics including age, sex, weight, chronic health data and baseline biochemistry. The proportion of patients with eGFR < 60ml/min/m^2^ and baseline creatinine will be presented. Other factors that may influence the primary outcome are also included such as medications, left ventricular ejection fraction and operative data.

### The ICU interventions provided to both study arms will be shown in

Table. These interventions will be divided into cardiovascular supports, fluid and diuretic therapy. Unless otherwise specified, interventions will be measured from the time of ICU admission following surgery to the time of ICU discharge.

**For the above tables, categorical data will be summarised as numbers and percentages, while continuous data will be displayed as means with standard deviations for normally distributed data or medians with inter-quartile ranges for skewed data. For**

Table, p-values will be generated using Fisher’s exact test for categorical data, Student’s t-test for normally distributed continuous data and the Mann-Whitney U test for skewed continuous data.

## ANALYSIS

### Analysis methods

**The analysis of the primary, secondary and tertiary outcomes will be presented in** Table.

### Main analysis of the primary outcome

The number and percentage of participants meeting the primary outcome after randomisation will be presented for each study arm. The primary effect estimate will be the relative risk of in-hospital acute kidney injury and will be reported with a 95% confidence interval. Comparison between treatment groups will be performed, unadjusted, using the χ^2^ test. A model adjusting for baseline eGFR and study site will also be reported. Odds ratio and 95% confidence interval will also be calculated. The primary outcome will also be analysed in the subgroups of patients with estimated glomerular filtration rates over and under 60ml/min/1.73m^2^.

### Analysis of secondary and tertiary outcomes

Outcomes with continuous dependent variables (ICU length of stay, hospital length of stay, duration of mechanical ventilation, serum albumin at 24 hours and net fluid balance at 48 hours post randomisation) will be presented as means with standard deviations or medians with inter-quartile ranges as appropriate. Comparison will be performed using student’s t-test for normally distributed data or the Mann-Whitney U test for skewed data. These outcomes will be presented as mean or median differences with 95% confidence intervals. Duration variables (ICU length of stay, hospital length of stay and duration of mechanical ventilation) will be calculated from the index ICU admission date and time and will be analysed using risk regression to account for competing risk of early death for all participants and the subset of survivors.

Major Adverse Kidney Events at day 28 (MAKE-28) will be defined as a composite outcome of hospital mortality up to day 28, or in hospital AKI stage II and III including dialysis up to day 28. MAKE-28, hospital mortality, in-hospital AKI stages 2 or 3 and in-hospital arrythmia will be presented as the number and percentage of participants meeting the outcome and compared using the χ^2^ test. Results will be reported as a relative risk with 95% confidence interval. Additionally, time to event analysis of hospital mortality up to day-28 will be analysed with a corresponding Kaplan-Meier curve. MAKE-28 will also be analysed in the subgroups of patients with estimated glomerular filtration rates over and under 60ml/min/1.73m^2^. A model adjusting for baseline eGFR and study site will also be reported for secondary and tertiary outcomes.

### Sub-group analyses

Logistic regression will be used to generate sub-group analyses of the primary outcome. Odds ratios with 95% confidence intervals will be presented in a forest plot for the sub-groups identified below:

- Baseline eGFR >60 vs <60 ml/min/1.73m^2^
- Patient age, above vs below median value
- Diabetes status, yes vs no
- Preoperative use of angiotensin receptor blocker or angiotensin converting enzyme inhibitor, yes vs no
- Sex, male vs female
- Cardiopulmonary bypass time >120 minutes vs <120 minutes
- Left ventricular function, moderate to severe dysfunction vs normal
- Site and region, Australian sites vs Italian site

### Sensitivity analyses

The following sensitivity analyses will be performed:

- The main analysis of the primary outcome will also be repeated using a per-protocol population should the number of patients with a major protocol deviation, defined as not receiving 20% albumin in the intervention group or receiving 20% albumin in the standard care group in the first 24h after randomisation, exceed 5%.
- To assess the impact of any albumin on the primary outcome, the main analysis will be repeated on a population that excludes standard care patients who received ≥ 500ml of normal serum albumin by the end of day 2.

### Missing data

Given the entire data set is collected during the hospital admission, the amount of missing data is expected to be low. However, should missing data exceed 5% it will be assumed to be missing at random and multiple imputations will be used. Ten imputed models will be generated and adjusted for relevant baseline characteristics with the pooled dataset used for analysis.

Analyses will be performed using Stata version 16 (StataCorp, College Station, TX) and R software (R Core Team, 2023, www.R-project.org).

To further inform the impact of 20% albumin administration, we will perform a Bayesian analysis of the primary outcome. A range of priors, including a non-informative prior, will be used to determine posterior probabilities that the infusion of 20% albumin reduced the incidence of in-hospital acute kidney injury following high-risk cardiac surgery. This analysis will be presented in a separate publication.

### Data extraction and monitoring

Data will be collected contemporaneously during the patient stay in hospital into the study database. Central risk-based monitoring will be conducted to ensure patient safety, consent compliance and completion of baseline data, all outcome data on 100% of patients.

## ETHICS AND DISSEMINATION

The Monash Health Lead Human Research and Ethics Committee approved the trial for all Australian sites under the National Mutual Acceptance scheme. Every participating site approved the trial through Site Specific Approval and Governance. Ethics approval from the Italian Medicine agency was obtained for the conduct of the trial in Italy. Patients who meet all inclusion criteria will be screened for eligibility. Whenever possible, consent will be obtained from participants prior to surgery. If this is not feasible, consent will be sought from the participant or their surrogate decision-maker soon after arrival to intensive care and before randomization. The main trial results will be presented at key national and international discipline related meetings. The main manuscript will be published with open access in a major peer reviewed general medical or specialty journal, if not simultaneous with presentation at a chosen meeting, soon after first public presentation.

### Trial registration number

This trial was registered with the Australian New Zealand Clinical Trials Registry - ACTRN12619001355167

## ACKNOWLEDGEMENTS

The original protocol for this trial was published in Trials in 2021 [17], and presented at the Australian and New Zealand Intensive Care Society Clinical Trials Group Annual Meeting in 2023.

We thank and acknowledge the support of the ALBICS-AKI investigators, without whom this trial could not take place: Claire Harington, Lewis Raymond, Peter Grant – Prince of Wales, and Prince of Wales Private Hospital, Randwick, New South Wales. Lisa Dougherty – Cabrini Health, Malvern, Victoria. Akinori Maeda, Atthapong Phongphithakchai, Sofia Spano – Austin Hospital, Melbourne, Victoria. David Bleetman, Phoebe Darlison, Jennifer Holmes, Melissa King – St Vincent’s Hospital Melbourne, Fitzroy, Victoria. Joanne McIntyre – Department of ICU, Flinders Medical Centre, Bedford Park, South Australia; All in Australia. Vanessa Bottari and Francesco Albano – Careggi University Hospital, Florence, Italy.

## AUTHORS’ CONTRIBUTIONS

YS is the chief principal investigator. Authors MB, AP, WA and YS contributed to the conception and design of this study. AP, VS, RB, AB, RM, SB and YS are principal investigators at their respective sites. All authors contributed to and reviewed the manuscript. All authors form the trial steering committee. All authors read and approved the final manuscript.

## DATA AVAILABILITY STATEMENT

Data sharing is not applicable to this article as no datasets were generated or analysed for the creation of this statistical analysis plan and updated protocol.

The datasets and statistical code for the completed study will be available according to Monash University data sharing protocols. The authors have no contractual agreements to disclose that would limit such access.

## TRANSPARENCY STATEMENT

All authors affirm that this manuscript is an honest, accurate, and transparent account of the study being reported; that no important aspects of the study have been omitted; and that any discrepancies from the study as planned and registered have been explained.

